# *HBA* Copy Number and Kidney Disease Risk among Black Americans: a Longitudinal Cohort Study

**DOI:** 10.1101/2021.04.01.21254397

**Authors:** A. Parker Ruhl, Neal Jeffries, Yu Yang, Rakhi P. Naik, Amit Patki, Lydia H. Pecker, Bryan T. Mott, Neil A. Zakai, Cheryl A. Winkler, Jeffrey B. Kopp, Leslie A. Lange, Marguerite R. Irvin, Orlando M. Gutierrez, Mary Cushman, Hans C. Ackerman

## Abstract

**Background:** Alpha globin gene (*HBA)* copy number is variable among people of African descent. *HBA* limits endothelial nitric oxide signaling and variation in gene copy number could modify kidney disease risk in this population.

**Objective:** To examine the association of *HBA* copy number with chronic kidney disease (CKD) and end-stage kidney disease (ESKD). We hypothesized that higher *HBA* copy number would be associated with greater CKD prevalence and ESKD incidence.

**Design:** Prospective, longitudinal cohort from the REasons for Geographic and Racial Differences in Stroke (REGARDS) study which enrolled participants from 2003 through 2007 and conducted follow-up through June 2014. Data were analyzed from January 2018 through January 2021.

**Setting:** Community-dwelling participants enrolled throughout the contiguous United States

**Participants:** Black Americans age 45 years and older

**Measurements:** *HBA* copy number was measured using droplet-digital PCR on genomic DNA; copy number ranged from 2 to ≥ 5 copies. The prevalence ratio (PR) of CKD and relative risk (RR) of incident reduced estimated glomerular filtration rate (eGFR) were calculated using modified Poisson multivariable regression employing a log-linear effect of *HBA* allele count. The hazard ratio (HR) of incident ESKD was calculated using Cox proportional hazards multivariable regression.

**Results:** Among 9,918 participants, *HBA* gene copy number frequencies were 4%, 28%, 67%, and 1% for 2, 3, 4, and ≥5 copies, respectively. After adjusting for demographic, clinical, and genetic risk factors, each additional copy of *HBA* was associated with 14% greater prevalence of CKD (PR = 1.14, 95% CI 1.07 to 1.21; *P* < 0.0001). While there was no significant association with incident reduced eGFR (RR = 1.06, 95% CI 0.94 to 1.18; p = 0.36), the hazard of incident ESKD was 28% higher for each additional copy of *HBA* (HR = 1.28, 95% CI 1.01 to 1.61; *P* = 0.04).

**Limitations:** This study did not identify the mechanism by which *HBA* copy number modifies kidney disease risk. This study focused on Black Americans, a population with a high frequency of the 3.7 kb gene deletion; it is unknown whether *HBA* modifies kidney disease risk in other populations with the 3.7 kb deletion.

**Conclusions:** Increasing *HBA* copy number was associated with greater prevalent CKD and incident ESKD in a national longitudinal study of Black Americans.

**Funding Source:** National Institutes of Health

## INTRODUCTION

Black Americans develop kidney disease at a younger age than other Americans and are three times more likely to develop end-stage kidney disease (ESKD), even after accounting for socioeconomic factors and comorbid medical conditions.^1–6^ DNA sequence variants that increase the risk of kidney disease, such as those in *HBB* (sickle cell trait)^7,8^ and *APOL1*^9–11^ are more common among Black Americans, yet only partly explain the racial disparity in kidney disease. The evaluation of genetic risk factors for diseases common in minority populations in the United States remains a high priority. However, Blacks and other minority populations are underrepresented in many large-scale longitudinal US population studies hindering genetic investigations.^12,13^

Basic science^14–17^ and clinical studies^18,19^ have identified nitric oxide signaling as important in protection against and recovery from ischemic or oxidative injury to the kidney; however, to date there have been few clear associations with genetic variants in nitric oxide signaling pathways in the kidney.^20–23^ Alpha globin, encoded by the tandem duplicated *HBA1* and *HBA2* genes on human chromosome 16, has been proposed to function as a regulator of nitric oxide signaling in the endothelial cells of small resistance arteries.^24^ Genetic or pharmacologic disruption of alpha globin or its stabilizing protein leads to enhanced nitric oxide signaling and decreased vasoconstriction in response to alpha-adrenergic stimuli,^25–27^ potentially conferring protection against kidney injury. The genes encoding alpha globin are highly polymorphic worldwide and a 3.7 kilobase insertion/deletion variant is common among Black Americans. This deletion spans parts of the *HBA2* and *HBA1* genes and reduces the functional gene copy number by one; leading to a decrease in *HBA* gene expression in red cell precursors.^28^ Due to the unique challenges of genotyping this structural variant, it has been omitted from genome-wide association studies. We hypothesized that higher *HBA* copy number would be associated with greater risk of prevalent chronic kidney disease (CKD), incident reduced eGFR, and incident ESKD among Black Americans.

## METHODS

### Study Design

The REasons for Geographic and Racial Differences in Stroke (REGARDS) study is a longitudinal cohort study designed to determine the reasons for racial disparities in stroke and cognitive decline in Black and White Americans aged ≥45 years.^29^ REGARDS enrolled 30,239 community-dwelling participants from the 48 contiguous United States from 2003 to 2007. A total of 12,514 (41%) participants were Black, and 56% were from states considered to be in the stroke belt, where stroke incidence in the United States is highest. Exclusion criteria included self-reported race other than Black / African-American or White, residence in or on the waiting list for a nursing home, active cancer within the past year, or inability to communicate in English.

Baseline variables were collected via standardized computer-assisted telephone interview, self-administered questionnaire, and in-home physical examination.^29,30^ Trained personnel administered computer-assisted telephone interviews to collect study participant age, sex, region of residence, insurance status, education level, income, and reports of physician-diagnosed comorbid conditions. Trained personnel conducted in-home examinations to measure height, weight, and blood pressure (BP); collect blood and urine specimens; and review medication bottles. Baseline measurements were repeated in some participants during a second in-home visit conducted a median (25^th^, 75^th^ percentile) of 9.5 (8.7, 9.9) years after baseline.

All participants provided oral and written informed consent. The REGARDS study was approved by participating center Institutional Review Boards. All self-reported Black participants consenting to genetic research were eligible to participate in this study. Participants were excluded if they had hemoglobin SS (n=5), SC (n=1) or CC (n=6), or had ESKD at enrollment (n=81) (Figure 1). This study followed the Strengthening the Reporting of Observational Studies in Epidemiology (STROBE) reporting guideline.

**Figure 1.**
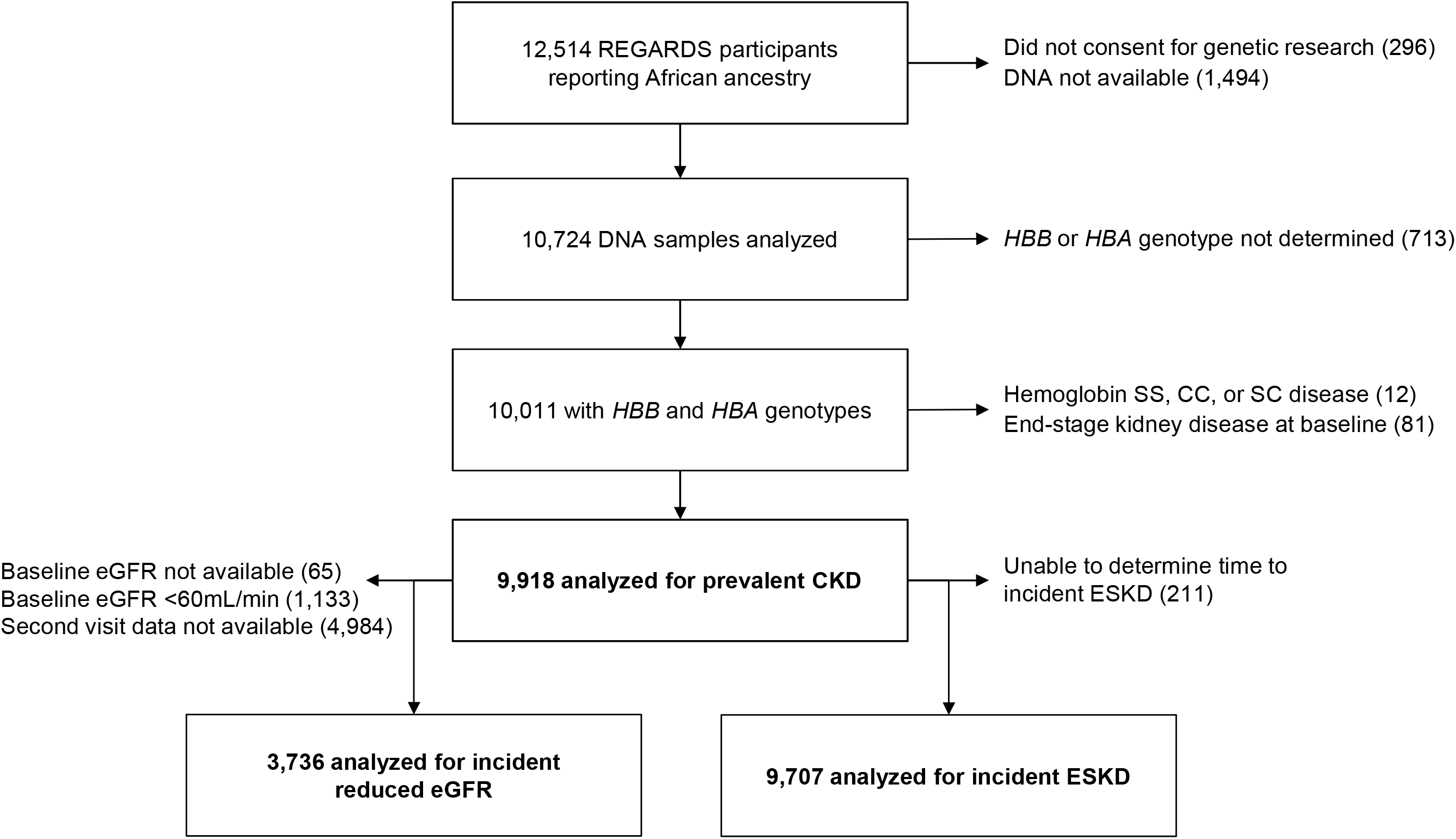
Cohort flow diagram. REGARDS= REasons for Geographic and Racial Differences in Stroke Study; *HBB*= beta globin gene; *HBA*= alpha globin gene; CKD= chronic kidney disease; ESKD= end-stage kidney disease; eGFR= estimated glomerular filtration rate

#### Main exposure variable

*HBA* copy number was evaluated as a numerical variable with values of 2, 3, 4, 5, or 6 as determined by droplet digital PCR (ddPCR) analysis of genomic DNA. The ddPCR copy number assay targeted a unique sequence within the 3.7 kb insertion/deletion polymorphism. We determined copy number of this target relative to a reference gene *EIF2C1*. Two-dimensional clusters of droplet counts for target and reference genes were manually gated using Quantasoft (Bio-Rad) per the manufacturer’s protocols. Droplet counts, copy number variant (CNV) values, and 95% CIs for CNV were extracted, visualized, and genotype was assigned using custom scripts in the R computing environment without user intervention (see Appendix).

#### Outcome measures

Prevalent CKD was defined by a urine albumin to creatinine ratio ≥30 mg/g or an estimated glomerular filtration rate (eGFR) < 60 mL/min/1.73m^2^ at baseline. The Chronic Kidney Disease Epidemiology Consortium equation was used to estimate GFR from serum creatinine concentrations. Incident reduced eGFR was defined by an eGFR < 60mL/min at the follow-up in-home visit and greater than 40% decline in eGFR from baseline, among those who had eGFR ≥ 60 mL/min at baseline. The incident reduced eGFR analysis was limited to those participants with available follow-up visit data (Figure 1). Incident ESKD was identified by linkage to United States Renal Data System (USRDS) data through June 30, 2014.

#### Covariates

Sickle cell trait and hemoglobin C genotypes were measured by a TaqMan single nucleotide polymorphism (SNP) genotyping assay (Applied Biosystems/Thermo Fisher Scientific).^8^ Apolipoprotein-L1 (*APOL1*) high-risk genotype was defined as the presence of two renal risk alleles compared to a reference of less than two high-risk alleles.^31,32^ Systolic (SBP) and diastolic blood pressure (DBP) were defined as the average of two measurements performed according to a standardized protocol.^33^ Hypertension was defined as one or more of the following: SBP ≥ 140 mmHg or DBP ≥ 90mmHg; self-reported current use of medication to control blood pressure; or two or more antihypertensive medications found on medication bottle review during the baseline in-home study visit. Hemoglobin, red blood cell (RBC) count and mean cell volume (MCV) values were measured with an automated hematology analyzer and mean cell hemoglobin (MCH), mean corpuscular hemoglobin concentration (MCHC), and red-cell distribution width-coefficient of variation (RDW-CV) calculated using standard formulae.^34^ Diabetes mellitus was defined by a fasting glucose level ≥126 mg/dL, a random glucose ≥200 mg/dL, or use of anti-diabetic medication. Body mass index was calculated from measured height and weight. Smoking status was self-reported and categorized as never, past, or current smoker. Region was defined as three geographic areas: stroke belt buckle (coastal plains of North Carolina, South Carolina, and Georgia), stroke belt (the rest of North Carolina, South Carolina, and Georgia and the entire states of Tennessee, Alabama, Mississippi, Arkansas, and Louisiana), and stroke nonbelt (the remaining contiguous US). Age, sex, race, health insurance (yes or no), highest education level obtained (less than high school, high school, some college, college or more), and annual income (≤$20K, $20-34K, $35-74K, ≥$75K) were self-reported.

### Statistical analysis

The analytic plan and outcome measure definitions were prespecified and approved by the REGARDS Cohort Study Executive Committee in December 2018.

#### Unadjusted analyses of participant characteristics by HBA copy number

For continuous measures of participant characteristics, medians and 25^th^ and 75^th^ percentiles were calculated by *HBA* copy number. Differences between copy number groups were assessed by Kruskal-Wallis non-parametric analysis of variance. Categorical variables were calculated as percentages within each category and differences were assessed by chi-squared tests of association.

#### Adjusted Analyses of HBA copy number and kidney disease outcome measures

The primary outcome measure was the presence or absence of prevalent CKD at baseline and the main exposure variable was *HBA* copy number. In addition, secondary outcome measures for incident reduced eGFR and incident ESKD were evaluated. A modified Poisson regression model with robust variance estimation was used to estimate the prevalence ratio (PR) and 95% confidence interval (CI) for CKD and the relative risk (RR) and 95% CI for incident reduced eGFR. ^35^ The models estimated the difference in log-transformed PR or RR for each additional *HBA* allele. Thirteen known risk factors for kidney disease were included in the multivariable regression models for prevalent CKD, incident reduced eGFR, and incident ESKD (see covariates section above). Incident ESKD hazard ratio (HR) and 95% CI was estimated with a multivariable Cox proportional-hazards model for the time between baseline and date of ESKD onset from USRDS.

#### Missing data considerations

Missing data for the outcome measures and explanatory variables were rare (< 0.5%) with some exceptions, e.g., complete blood count results (Table 1). A multiple imputation approach was employed in all multivariable regression models (see Appendix).^36^

**Table 1.** Clinical and demographic characteristics according to *HBA* copy number.

| Participants,<br>N (%) | *HBA* copy number |  |  |  |  | *P* value<sup>†</sup> |
| --- | --- | --- | --- | --- | --- | --- |
|  | All<br>Participants | 2 | 3 | 4 | ≥ 5* |  |
|  | 9918 (100) | 393 (4) | 2747 (28) | 6674 (67) | 104 (1) | NA |
| **Age, years** | 63.0<br>(57.0, 70.0) | 64.0<br>(58.0, 70.0) | 64.0<br>(57.0, 71.0) | 63.0<br>(57.0, 70.0) | 66.5<br>(61.0, 72.2) | 0.01 |
| **Female sex, N (%)** | 6100 (62) | 246 (63) | 1681 (61) | 4115 (62) | 58 (56) | 0.61 |
| **Body mass index,**<br>**kg/m<sup>2</sup>** | 29.7<br>(26.2, 34.4) | 30.3<br>(26.8, 31.2) | 29.9<br>(26.1, 34.4) | 29.7<br>(26.1, 30.7) | 30.4<br>(27.6, 30.9) | 0.22 |
| **Smoking status, N**<br>**(%)** |  |  |  |  |  | 0.55 |
| Never | 4478 (45) | 169 (43) | 1256 (46) | 3007 (45) | 46 (44) |  |
| Past | 3675 (37) | 160 (41) | 1007 (37) | 2464 (37) | 44 (42) |  |
| Current | 1720 (17) | 61 (16) | 470 (17) | 1175 (18) | 14 (13) |  |
| **Region, N (%)** |  |  |  |  |  | 0.06 |
| Non-Belt | 4936 (50) | 170 (43) | 1397 (51) | 3309 (50) | 60 (58) |  |
| Belt | 3286 (33) | 142 (36) | 881 (32) | 2235 (33) | 28 (27) |  |
| Buckle | 1696 (17) | 81 (21) | 469 (17) | 1130 (17) | 16 (15) |  |
| **Medically insured,**<br>**N (%)** | 8927 (90) | 355 (91) | 2489 (91) | 5987 (90) | 96 (92) | 0.36 |
| **Education level, N**<br>**(%)** |  |  |  |  |  | 0.29 |
| Less than high<br>school | 1939 (20) | 71 (18) | 547 (20) | 1299 (19) | 22 (21) |  |
| High school graduate | 2730 (28) | 111 (28) | 787 (29) | 1803 (27) | 29 (28) |  |
| Some college | 2660 (27) | 105 (27) | 756 (28) | 1770 (27) | 29 (28) |  |
| College graduate or more | 2577 (26) | 105 (27) | 655 (24) | 1793 (27) | 24 (23) |  |
| **Income, N (%)** |  |  |  |  |  | 0.12 |
| ≤ \$20K | 2654 (30) | 104 (30) | 746 (31) | 1773 (30) | 31 (32) | |
| \$20K - \$34K | 2611 (30) | 109 (31) | 725 (30) | 1744 (30) | 33 (34) | |
| \$35K - \$74K | 2538 (29) | 106 (30) | 718 (30) | 1688 (29) | 26 (27) | |
| ≥ \$75K | 914 (10) | 32 (9) | 214 (9) | 662 (11) | N < 11 | |
| **Hemoglobin, per g/dL** | 13.1<br>(12.2, 14.0) | 12.3<br>(11.6, 13.2) | 12.9<br>(12.1, 13.8) | 13.2<br>(12.4, 14.1) | 13.0<br>(12.0, 14.1) | <0.0001 |
| **RBC count, million cells/cmm** | 4.5<br>(4.2, 4.8) | 5.2<br>(4.7, 5.6) | 4.7<br>(4.3, 5.0) | 4.4<br>(4.1, 4.7) | 4.4<br>(4.0, 4.7) | <0.0001 |
| **MCV, fL** | 88.0<br>(84.0, 92.0) | 74.0<br>(72.0, 77.0) | 84.0<br>(82.0, 87.0) | 90.0<br>(87.0, 93.0) | 88.0<br>(86.0, 92.0) | <0.0001 |
| **MCH, pg** | 29.6<br>(27.9, 30.9) | 23.8<br>(22.8, 24.8) | 27.9<br>(26.9, 29.0) | 30.3<br>(29.2, 31.4) | 29.9<br>(29.1, 30.9) | <0.0001 |
| **MCHC, g/dL** | 33.4<br>(32.9, 33.9) | 32.1<br>(31.6, 32.6) | 33.0<br>(32.6, 33.5) | 33.7<br>(33.2, 34.1) | 33.7<br>(33.2, 34.0) | <0.0001 |
| **RDW-CV, %** | 14.0<br>(13.3, 14.8) | 15.0<br>(14.4, 15.9) | 14.2<br>(13.5, 15.0) | 13.8<br>(13.2, 14.6) | 13.6<br>(13.2, 14.2) | <0.0001 |
| **eGFR <60 mL/min/1.73 m<sup>2</sup>, N (%)** | 1133 (11) | 37 (9) | 300 (11) | 779 (12) | 17 (16) | 0.16 |
| **Urine albumin to creatinine ratio >30mg/g, N (%)** | 1794 (19) | 59 (16) | 498 (19) | 1214 (19) | 23 (24) | 0.23 |
| **Prevalent chronic kidney disease<sup>†</sup>, N (%)** | 2476 (26) | 80 (21) | 672 (25) | 1688 (26) | 36 (36) | 0.01 |
| **Hypertension, N (%)** | 7314 (77) | 298 (78) | 2032 (77) | 4900 (76) | 84 (83) | 0.31 |
| **Diabetes mellitus, N (%)** | 2850 (29) | 103 (26) | 818 (30) | 1900 (29) | 29 (28) | 0.41 |
| **Sickle cell trait, N (%)** | 743 (7) | 25 (6) | 211 (8) | 501 (8) | N < 11 | 0.73 |
| ***APOL1* high-risk, N (%)** | 1251 (13) | 60 (15) | 350 (13) | 830 (13) | 11 (11) | 0.38 |

*HBA*= alpha globin gene; N= number; K= thousand; RBC= red blood cell; MCV= mean corpuscular volume; MCH= mean corpuscular hemoglobin; MCHC=mean corpuscular hemoglobin concentration; RDW-CV= red cell distribution width-coefficient of variation; eGFR= estimated glomerular filtration rate; *APOL1*= apolipoprotein-L1. Values are median (25<sup>th</sup>, 75<sup>th</sup> percentile) except where otherwise indicated.
\*102 participants were found to have 5 *HBA* gene copies and 2 were found to have 6 *HBA* gene copies; <sup>†</sup>P values for tests of differences by *HBA* copy number generated from the chi-squared test for categorical variables and the Kruskal-Wallis non-parametric ANOVA test for continuous variables; <sup>‡</sup>Prevalent CKD was defined by a urine albumin to creatinine ratio $\geq 30$ mg/g or an estimated glomerular filtration rate (eGFR) < 60 mL/min/1.73m<sup>2</sup> at baseline.
Missing data are as follows: Smoking status (n=45, 0.5%); insured (n=12, 0.1%); education (n=12, 0.1%); income (n=1,201 refused, 12.1%); eGFR (n=65, 0.7%); urine albumin creatinine ratio (n=398, 4.0%); hemoglobin (n=3,193, 32.2%); RBC count (n=3,193, 32.2%); MCV (n=3,200, 32.3%); MCH (n=3,193, 32.2%); MCHC (n=3,193, 32.2%); RDW-CV (n=3,206, 32.3%); prevalent chronic kidney disease (n=367, 3.7%); hypertension (n=389, 3.9%); diabetes mellitus (n=51, 0.5%); body mass index (n=73, 0.7%); *APOL1* (n=64, 0.6%). Denominators for
percentages in the table are based on the number of non-missing responses. Due to rounding, percentages may not sum to 100%.

#### Sensitivity analyses and tests for interaction

To address the possibility of association by population stratification, we performed a sensitivity analysis including the first ten principal components of ancestry among 7,641 (77%) participants for whom Infinium Expanded Multi-Ethnic Genotyping Array data were available.

A pre-specified test for interaction between *HBA* copy number and SCT on the outcomes of prevalent CKD, incident reduced eGFR, and incident ESKD was performed for the fully adjusted multivariable models. Additionally, pre-specified tests for interaction between *HBA* copy number and each of the following variables, *APOL1*, age, sex, and hypertension, were performed on the outcome of prevalent CKD for the fully adjusted multivariable model.

#### Relationships between the structural variant and flanking SNPs

To exclude the possibility that the associations with kidney disease risk measured at the structural variant in the *HBA1* and *HBA2* loci were due to linkage disequilibrium with flanking SNPs, we analyzed SNP genotypes available on 8,841 participants from the Infinium Expanded Multi-Ethnic Genotyping Array. Pairwise linkage disequilibrium was calculated between the −3.7 kb structural variant and each of 13,549 SNPs in the first 1 Mb of chromosome 16 using the *r*^2^ statistic in the R (v 3.6.2) package genetics. Associations between individual SNPs and CKD prevalence were calculated using PLINK (v 1.9) in a regression model that included age, sex, and the first 4 prinicipal components of ancestry. The threshold for significance for a 1 Mb segment of the genome was estimated to be p < 1.5 × 10^−4^ based on the threshold for genome-wide significance(*p* < 5 × 10^−8^) divided by the fraction of the genome (1 Mb / 3,097 Mb) analyzed.^38^

#### Population preventable fraction

To estimate the fraction of kidney disease prevented by the protective genotypes of *HBA* copy numbers 2 or 3 among Black Americans, we calculated the population preventable fraction and the upper and lower bounds of the 95% CI for the fully adjusted models (further methods in the Appendix).

## RESULTS

### HBA copy number variation

*HBA* copy number was variable among the 9,918 participants who met inclusion and exclusion criteria (Figure 1): 393 participants (4%) had 2 *HBA* copies, 2,747 (28%) had 3 *HBA* copies, 6,674 (67%) had 4 *HBA* copies, 102 (1%) had 5 *HBA* copies, and 2 (<1%) participants had 6 *HBA* copies (Table 1). CKD prevalence increased with *HBA* copy number. It was 21% among those with 2 *HBA* copies; 25% among those with 3 copies; 26% among those with 4 copies; and 36% among those with ≥5 copies (p = 0.01; Table 1). Red blood cell parameters differed according to *HBA* copy number (p < 0.0001): a hypochromic, microcytic anemia was observed in those with 2 copies of *HBA*.

### HBA copy number and kidney disease outcome measures

#### Prevalent chronic kidney disease

A total of 2,476 (26%) participants had CKD at baseline. In an unadjusted analysis between *HBA* copy number and prevalent CKD, each additional copy of *HBA* was associated with a 9% greater risk of prevalent CKD (PR = 1.09 [95% CI 1.02, 1.15; p=0.009]). After adjusting for 13 CKD risk factors selected *a priori*, each additional copy of *HBA* was associated with a 14% greater prevalence of CKD (PR = 1.14 [95% CI 1.07 - 1.21; p< 0.0001]); Table 2 and Appendix Figure 1.

**Table 2.**
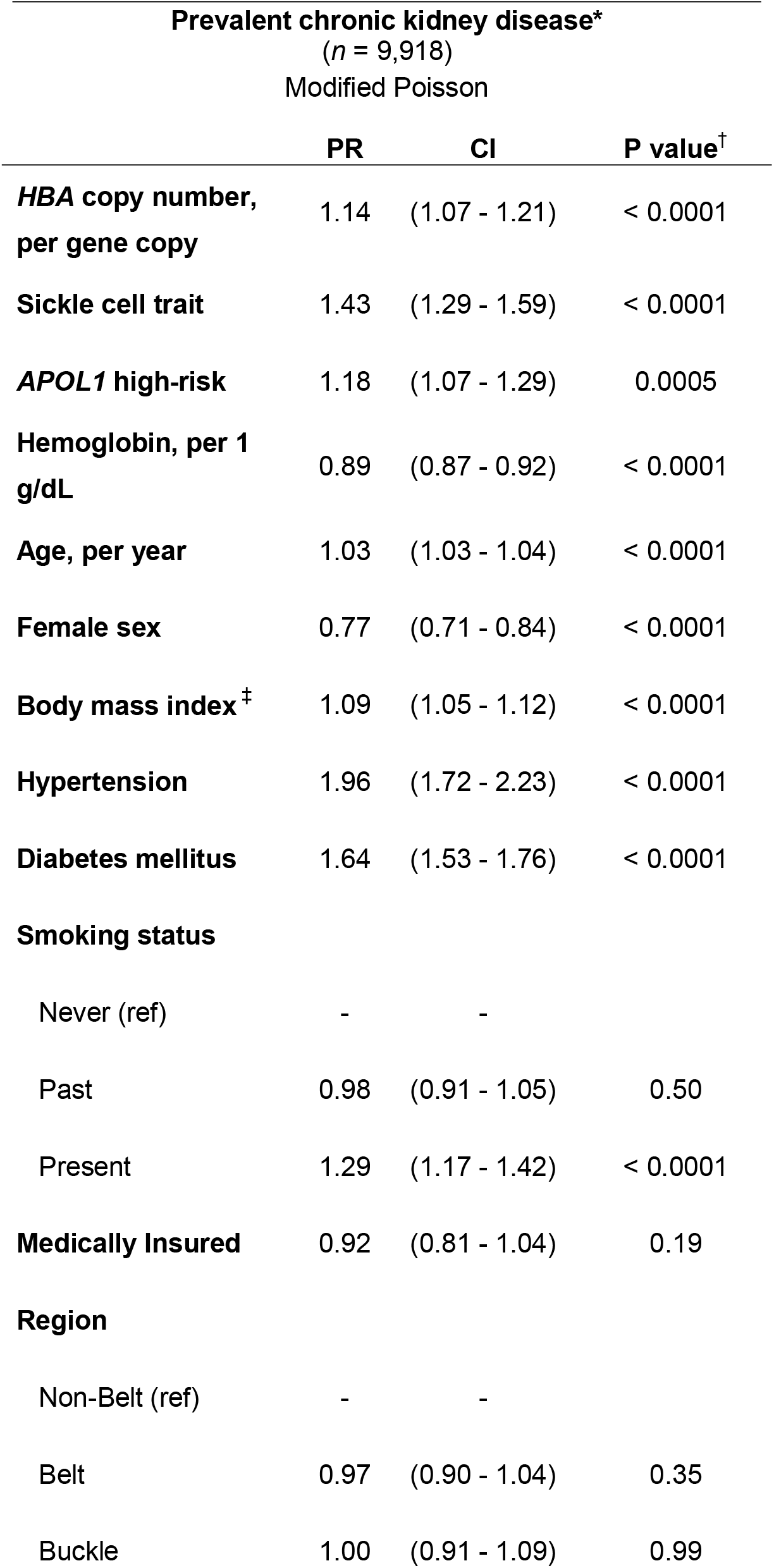
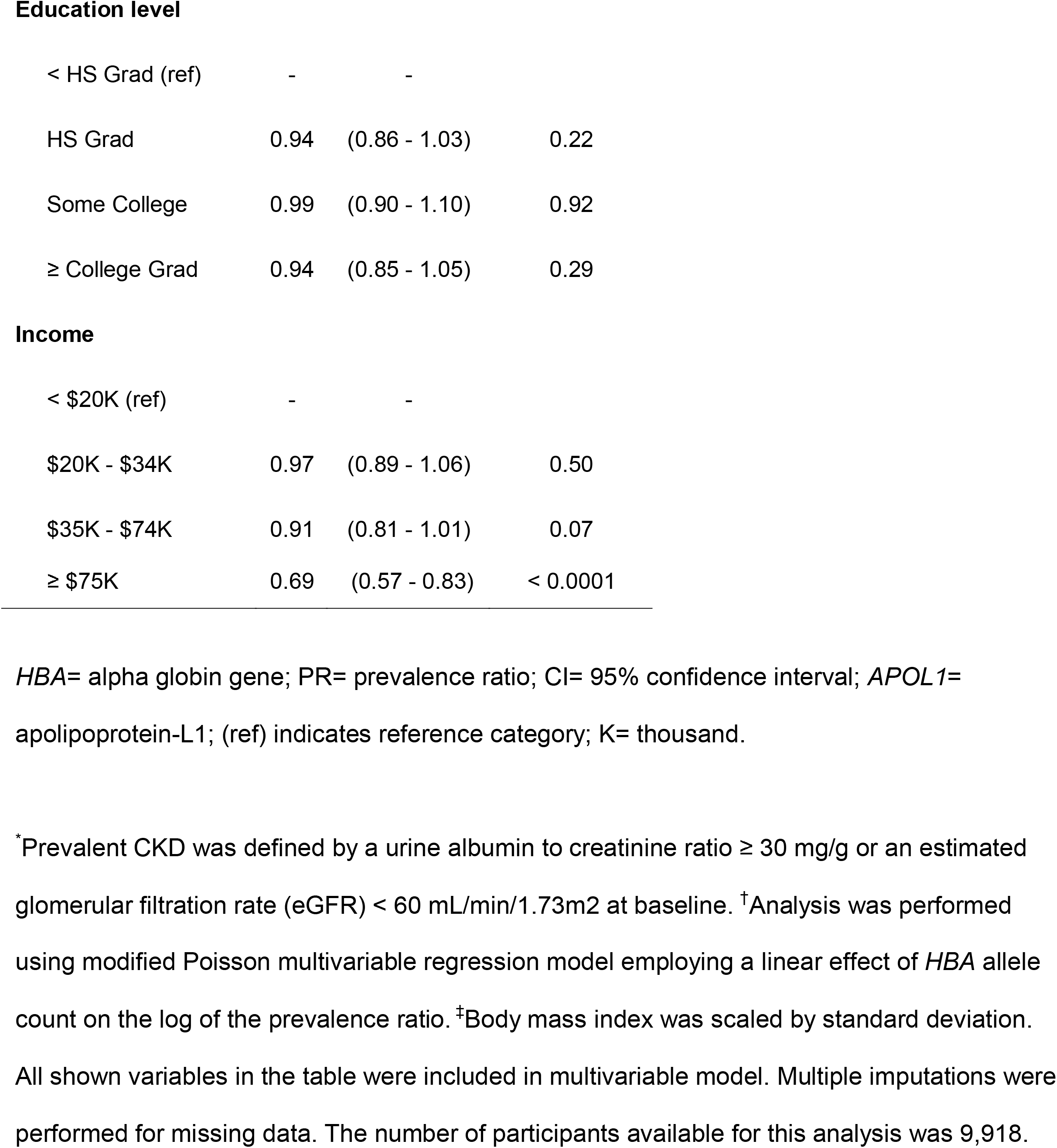
Association of *HBA* gene copy number with prevalent chronic kidney disease – analysis adjusted for all listed covariates

| Prevalent chronic kidney disease*<br>(*n* = 9,918)<br>Modified Poisson |  |  |  |
| --- | --- | --- | --- |
|  | PR | CI | P value<sup>†</sup> |
| ***HBA* copy number, per gene copy** | 1.14 | (1.07 - 1.21) | < 0.0001 |
| **Sickle cell trait** | 1.43 | (1.29 - 1.59) | < 0.0001 |
| ***APOL1* high-risk** | 1.18 | (1.07 - 1.29) | 0.0005 |
| **Hemoglobin, per 1 g/dL** | 0.89 | (0.87 - 0.92) | < 0.0001 |
| **Age, per year** | 1.03 | (1.03 - 1.04) | < 0.0001 |
| **Female sex** | 0.77 | (0.71 - 0.84) | < 0.0001 |
| **Body mass index<sup>‡</sup>** | 1.09 | (1.05 - 1.12) | < 0.0001 |
| **Hypertension** | 1.96 | (1.72 - 2.23) | < 0.0001 |
| **Diabetes mellitus** | 1.64 | (1.53 - 1.76) | < 0.0001 |
| **Smoking status** |  |  |  |
| Never (ref) | - | - |  |
| Past | 0.98 | (0.91 - 1.05) | 0.50 |
| Present | 1.29 | (1.17 - 1.42) | < 0.0001 |
| **Medically Insured** | 0.92 | (0.81 - 1.04) | 0.19 |
| **Region** |  |  |  |
| Non-Belt (ref) | - | - |  |
| Belt | 0.97 | (0.90 - 1.04) | 0.35 |
| Buckle | 1.00 | (0.91 - 1.09) | 0.99 |
| **Education level** |  |  |  |
| < HS Grad (ref) | - | - |  |
| HS Grad | 0.94 | (0.86 - 1.03) | 0.22 |
| Some College | 0.99 | (0.90 - 1.10) | 0.92 |
| ≥ College Grad | 0.94 | (0.85 - 1.05) | 0.29 |
| **Income** |  |  |  |
| < \$20K (ref) | - | - | |
| \$20K - \$34K | 0.97 | (0.89 - 1.06) | 0.50 |
| \$35K - \$74K | 0.91 | (0.81 - 1.01) | 0.07 |
| ≥ \$75K | 0.69 | (0.57 - 0.83) | < 0.0001 |

*HBA*= alpha globin gene; PR= prevalence ratio; CI= 95% confidence interval; *APOL1*= apolipoprotein-L1; (ref) indicates reference category; K= thousand.
\*Prevalent CKD was defined by a urine albumin to creatinine ratio ≥ 30 mg/g or an estimated glomerular filtration rate (eGFR) < 60 mL/min/1.73m<sup>2</sup> at baseline. <sup>†</sup>Analysis was performed using modified Poisson multivariable regression model employing a linear effect of *HBA* allele count on the log of the prevalence ratio. <sup>‡</sup>Body mass index was scaled by standard deviation. All shown variables in the table were included in multivariable model. Multiple imputations were performed for missing data. The number of participants available for this analysis was 9,918.

#### Incident reduced eGFR

Data were available for 3,736 participants who were evaluated at a second in-home visit (Figure 1). A total of 656 (17.6%) developed incident reduced eGFR over a median (25^th^, 75^th^ percentile) of 9.5 (8.7, 9.9) years. In an unadjusted analysis there was no association between *HBA* copy number and incident reduced eGFR (RR = 1.01 [95% CI 0.90 – 1.14; p=0.82]). After adjusting for relevant risk factors, there was no association between *HBA* copy number and incident reduced eGFR (RR = 1.06 [95% CI 0.94 - 1.18; p=0.36] Table 3).

**Table 3.**
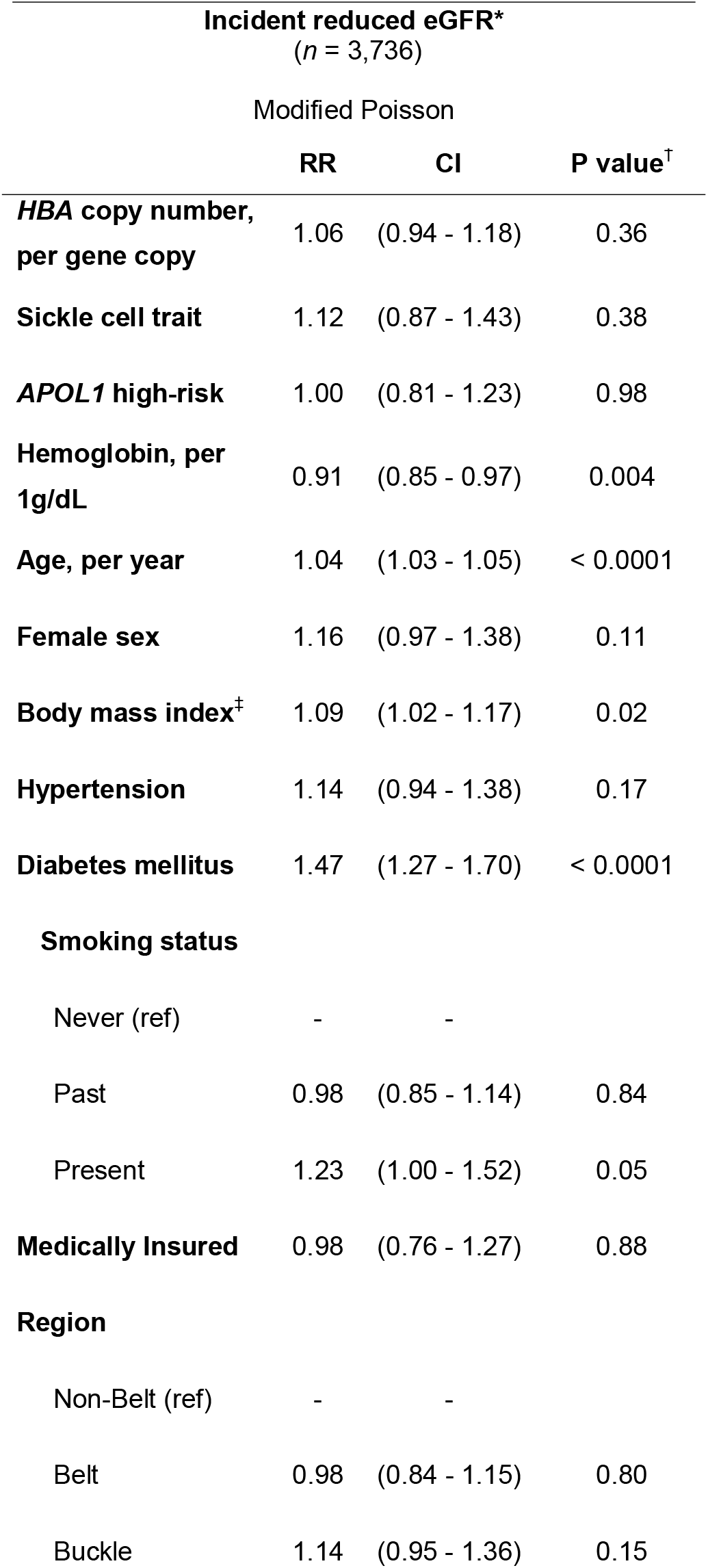
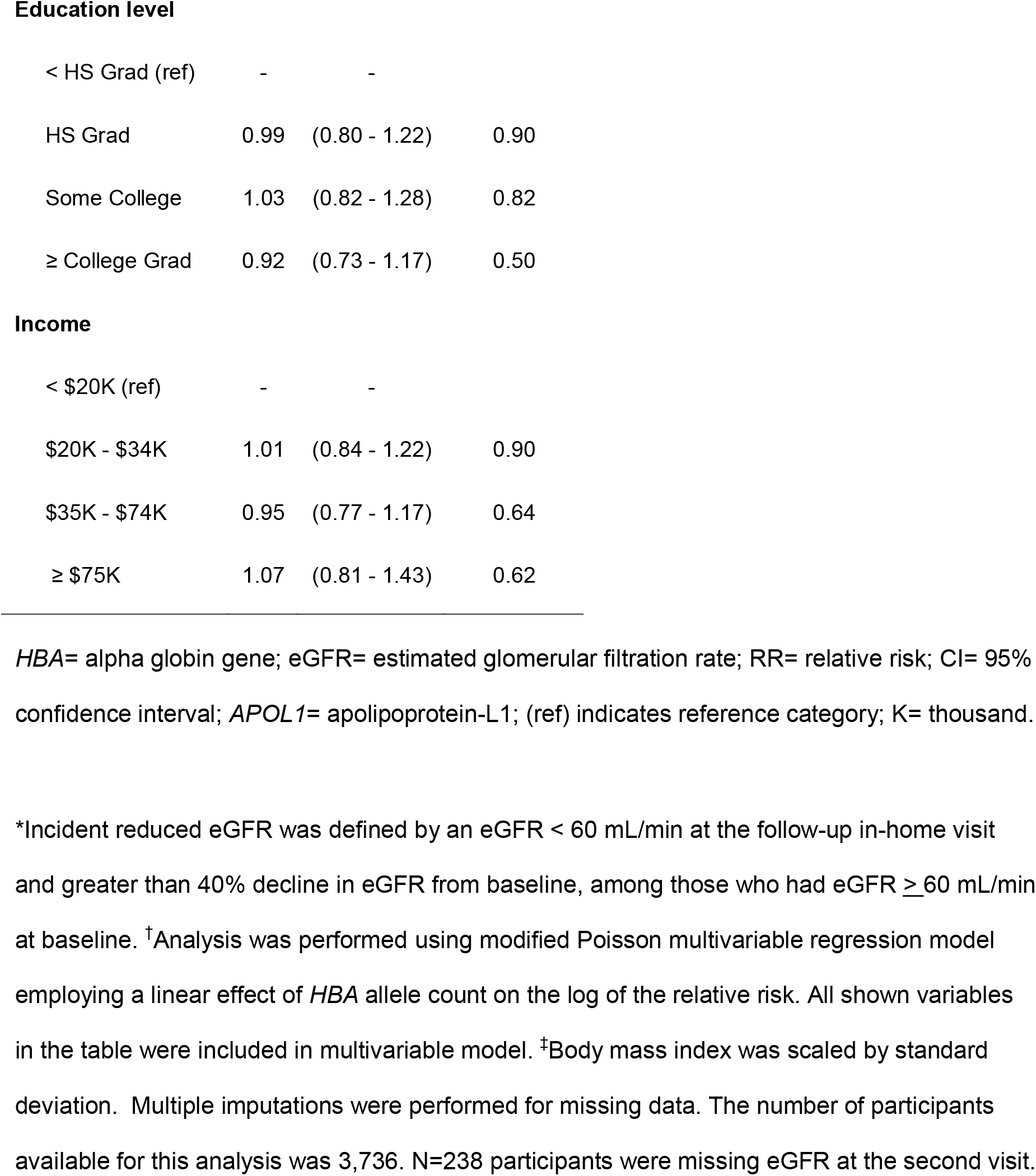
Association of *HBA* gene copy number with incident reduced eGFR – analysis adjusted for all listed covariates

| Incident reduced eGFR*<br>(*n* = 3,736) |  |  |  |
| --- | --- | --- | --- |
| Modified Poisson |  |  |  |
|  | RR | CI | P value<sup>†</sup> |
| ***HBA* copy number, per gene copy** | 1.06 | (0.94 - 1.18) | 0.36 |
| **Sickle cell trait** | 1.12 | (0.87 - 1.43) | 0.38 |
| ***APOL1* high-risk** | 1.00 | (0.81 - 1.23) | 0.98 |
| **Hemoglobin, per 1g/dL** | 0.91 | (0.85 - 0.97) | 0.004 |
| **Age, per year** | 1.04 | (1.03 - 1.05) | < 0.0001 |
| **Female sex** | 1.16 | (0.97 - 1.38) | 0.11 |
| **Body mass index<sup>‡</sup>** | 1.09 | (1.02 - 1.17) | 0.02 |
| **Hypertension** | 1.14 | (0.94 - 1.38) | 0.17 |
| **Diabetes mellitus** | 1.47 | (1.27 - 1.70) | < 0.0001 |
| **Smoking status** |  |  |  |
| Never (ref) | - | - |  |
| Past | 0.98 | (0.85 - 1.14) | 0.84 |
| Present | 1.23 | (1.00 - 1.52) | 0.05 |
| **Medically Insured** | 0.98 | (0.76 - 1.27) | 0.88 |
| **Region** |  |  |  |
| Non-Belt (ref) | - | - |  |
| Belt | 0.98 | (0.84 - 1.15) | 0.80 |
| Buckle | 1.14 | (0.95 - 1.36) | 0.15 |
| **Education level** |  |  |  |
| < HS Grad (ref) | - | - |  |
| HS Grad | 0.99 | (0.80 - 1.22) | 0.90 |
| Some College | 1.03 | (0.82 - 1.28) | 0.82 |
| ≥ College Grad | 0.92 | (0.73 - 1.17) | 0.50 |
| **Income** |  |  |  |
| < \$20K (ref) | - | - | |
| \$20K - \$34K | 1.01 | (0.84 - 1.22) | 0.90 |
| \$35K - \$74K | 0.95 | (0.77 - 1.17) | 0.64 |
| ≥ \$75K | 1.07 | (0.81 - 1.43) | 0.62 |

*HBA*= alpha globin gene; eGFR= estimated glomerular filtration rate; RR= relative risk; CI= 95% confidence interval; *APOL1*= apolipoprotein-L1; (ref) indicates reference category; K= thousand.
\*Incident reduced eGFR was defined by an eGFR < 60 mL/min at the follow-up in-home visit and greater than 40% decline in eGFR from baseline, among those who had eGFR ≥ 60 mL/min at baseline. <sup>†</sup>Analysis was performed using modified Poisson multivariable regression model employing a linear effect of *HBA* allele count on the log of the relative risk. All shown variables in the table were included in multivariable model. <sup>‡</sup>Body mass index was scaled by standard deviation. Multiple imputations were performed for missing data. The number of participants available for this analysis was 3,736. N=238 participants were missing eGFR at the second visit.

#### Incident end-stage kidney disease

Out of 9,707 eligible participants for the ESKD analysis, 234 participants developed ESKD over a median (25^th^, 75^th^ percentile) of 10.1 (5.5, 12.5) years of follow-up. In an unadjusted analysis there was no significant association between *HBA* copy number and the hazard of incident ESKD (HR = 1.02 [95% CI 0.82 – 1.28; p=0.84]). In contrast, an analysis adjusted for 13 risk factors selected *a priori* found that each additional copy of *HBA* was associated with a 28% increase in the hazard of incident ESKD (HR = 1.28 [95% CI 1.01 - 1.61; p=0.04] Table 4).

**Table 4.**
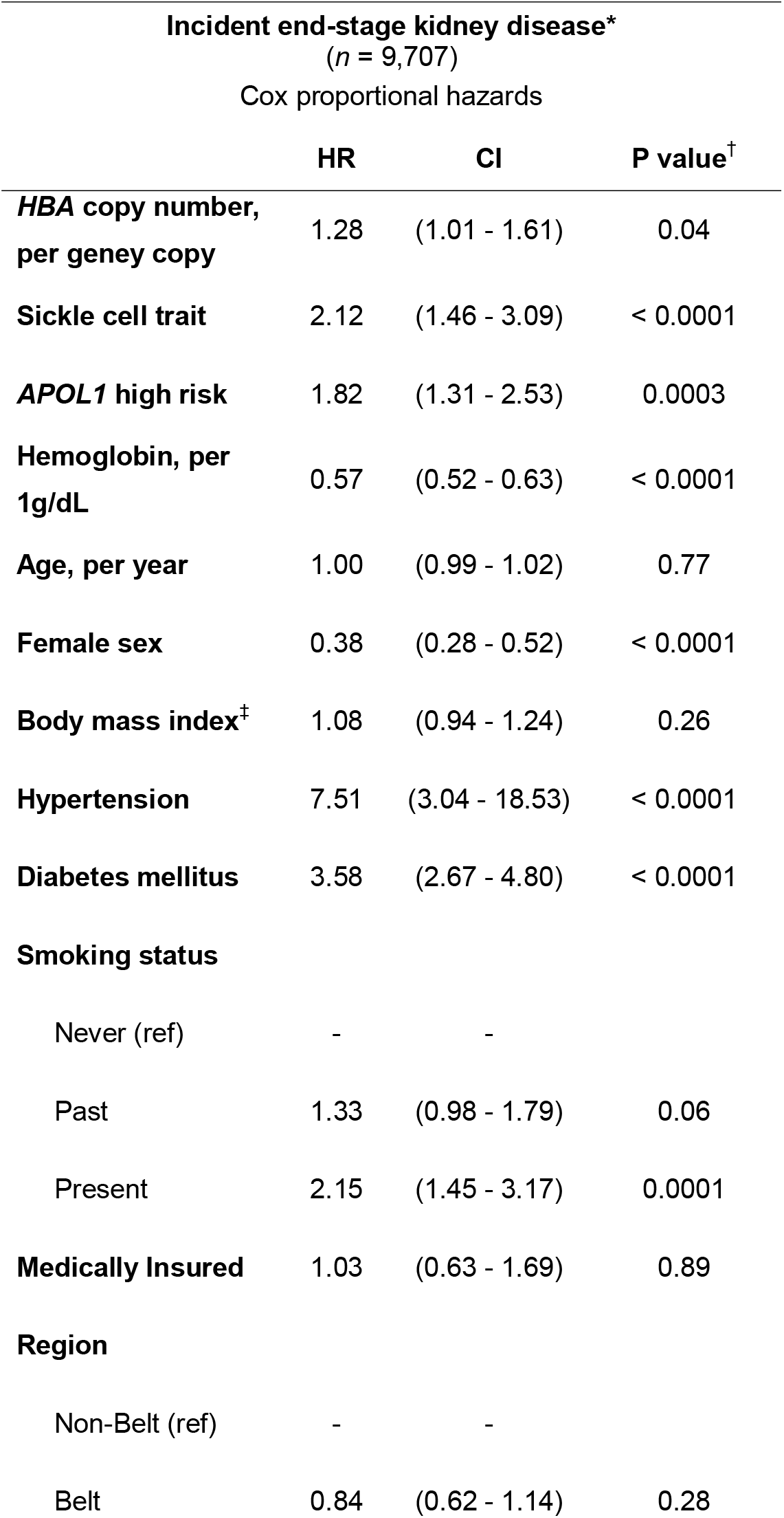
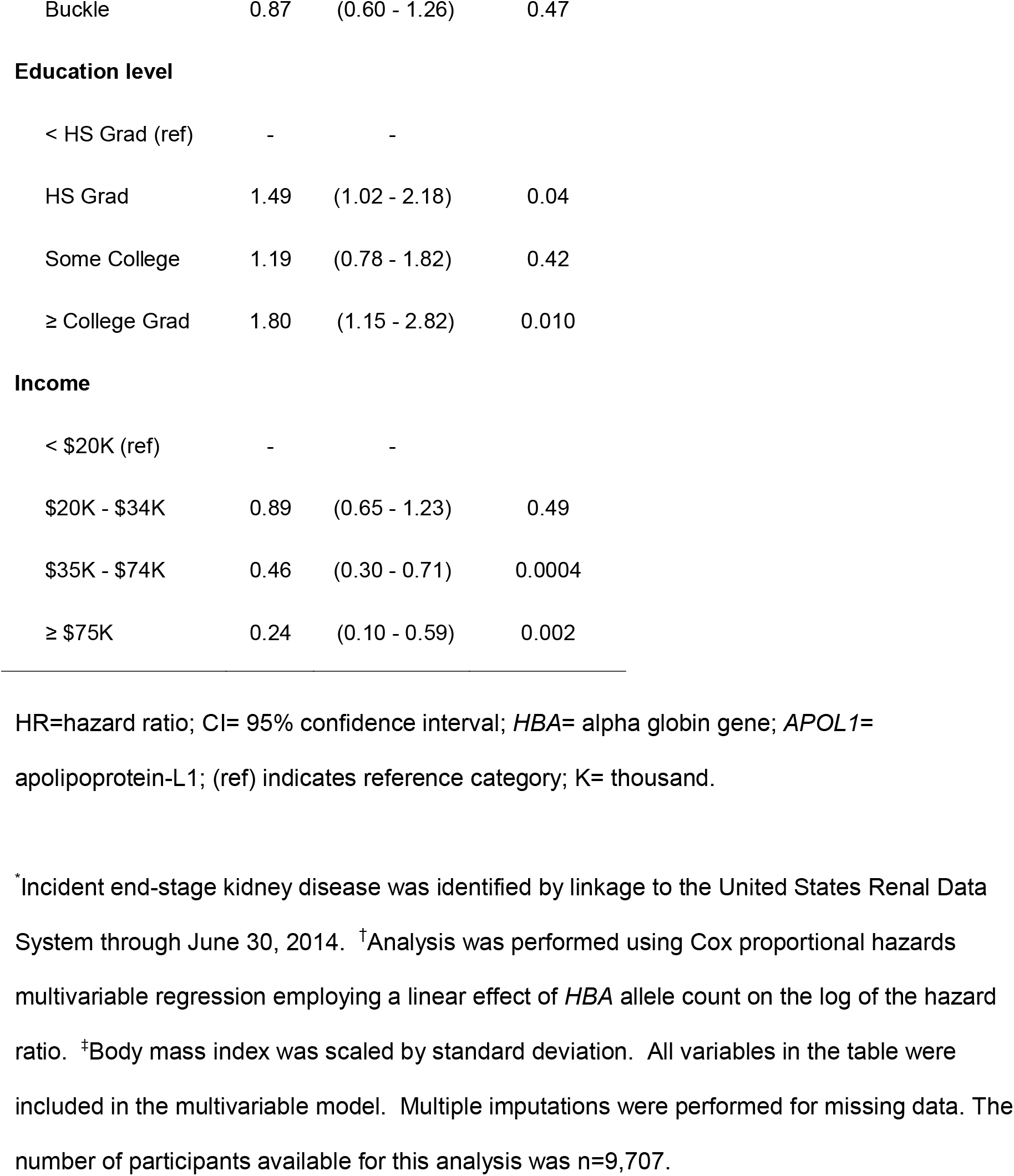
Association of *HBA* gene copy number with incident end-stage kidney disease – analysis adjusted for all listed covariates

| Incident end-stage kidney disease*<br>(*n* = 9,707)<br>Cox proportional hazards |  |  |  |
| --- | --- | --- | --- |
|  | HR | CI | P value<sup>†</sup> |
| ***HBA* copy number, per gene copy** | 1.28 | (1.01 - 1.61) | 0.04 |
| **Sickle cell trait** | 2.12 | (1.46 - 3.09) | < 0.0001 |
| ***APOL1* high risk** | 1.82 | (1.31 - 2.53) | 0.0003 |
| **Hemoglobin, per 1g/dL** | 0.57 | (0.52 - 0.63) | < 0.0001 |
| **Age, per year** | 1.00 | (0.99 - 1.02) | 0.77 |
| **Female sex** | 0.38 | (0.28 - 0.52) | < 0.0001 |
| **Body mass index<sup>‡</sup>** | 1.08 | (0.94 - 1.24) | 0.26 |
| **Hypertension** | 7.51 | (3.04 - 18.53) | < 0.0001 |
| **Diabetes mellitus** | 3.58 | (2.67 - 4.80) | < 0.0001 |
| **Smoking status** |  |  |  |
| Never (ref) | - | - |  |
| Past | 1.33 | (0.98 - 1.79) | 0.06 |
| Present | 2.15 | (1.45 - 3.17) | 0.0001 |
| **Medically Insured** | 1.03 | (0.63 - 1.69) | 0.89 |
| **Region** |  |  |  |
| Non-Belt (ref) | - | - |  |
| Belt | 0.84 | (0.62 - 1.14) | 0.28 |
| Buckle | 0.87 | (0.60 - 1.26) | 0.47 |
| Education level |  |  |  |
| < HS Grad (ref) | - | - |  |
| HS Grad | 1.49 | (1.02 - 2.18) | 0.04 |
| Some College | 1.19 | (0.78 - 1.82) | 0.42 |
| ≥ College Grad | 1.80 | (1.15 - 2.82) | 0.010 |
| Income |  |  |  |
| < \$20K (ref) | - | - | |
| \$20K - \$34K | 0.89 | (0.65 - 1.23) | 0.49 |
| \$35K - \$74K | 0.46 | (0.30 - 0.71) | 0.0004 |
| ≥ \$75K | 0.24 | (0.10 - 0.59) | 0.002 |

HR=hazard ratio; CI= 95% confidence interval; *HBA*= alpha globin gene; *APOL1*= apolipoprotein-L1; (ref) indicates reference category; K= thousand.
\*Incident end-stage kidney disease was identified by linkage to the United States Renal Data System through June 30, 2014. †Analysis was performed using Cox proportional hazards multivariable regression employing a linear effect of *HBA* allele count on the log of the hazard ratio. ‡Body mass index was scaled by standard deviation. All variables in the table were included in the multivariable model. Multiple imputations were performed for missing data. The number of participants available for this analysis was n=9,707.

#### Sensitivity analyses and tests of interaction

The point estimates of the associations between *HBA* copy number and the outcomes of prevalent CKD, incident reduced eGFR, and incident ESKD were not substantially changed and remained significant for both prevalent CKD and incident ESKD after adjustment for the first ten principal components of ancestry (Appendix Table 1). There was no interaction between *HBA* copy number and SCT for the outcomes of prevalent CKD, incident reduced eGFR, or incident ESKD (Appendix Table 2), nor was there an interaction between *HBA* copy number and each of age, sex, hypertension, or *APOL1* on the outcome of prevalent CKD (Appendix Table 3). The p-value of each cross-product interaction term was ≥ 0.20 in the fully-adjusted models.

#### Relationships between the HBA structural variant and nearby sequence variants

To assess whether associations with *HBA* copy number could be attributed to sequence variants flanking the *HBA1* and *HBA2* genes, we examined pairwise linkage disequilibrium between the −3.7 kb structural variant and 13,549 single nucleotide polymorphisms (SNPs) within the first 1 Mb of the chromosome. Pairwise linkage disequilibrium was weak between the structural variant and each SNP; the maximum *r*^*2*^ value was 0.25 and most SNPs had *r*^*2*^ values close to zero (Appendix Figure 2 A,B). Each SNP was tested for association with prevalent CKD. None of the SNPs reached the threshold for significance for a 1 Mb genome region (Appendix Figures 2 C,D), suggesting that the association between the structural variant and kidney disease risk was not due to linkage disequilibrium with SNPs in flanking gene regions.

#### Population preventable fraction

To assess the fraction of kidney disease prevented by the protective 2- and 3-copy *HBA* genotypes, we estimated the population preventable fraction. The 2- and 3-copy *HBA* genotypes prevent 4.3% (95% CI 2.1 - 6.5) of prevalent CKD and 11.4% (95% CI: 2.2 - 20.6) of incident ESKD among Black Americans (see Appendix).

## DISCUSSION

In this national longitudinal cohort study of Black Americans, each additional *HBA* copy was associated with a 14% greater prevalence of CKD and a 28% greater risk of incident ESKD after adjustment for established genetic, biomedical, demographic, and social risk factors. Kidney disease risk increased with higher *HBA* copy number, suggesting that the level of *HBA* gene expression, rather than a specific allele, was responsible for this genetic association. These findings support the hypothesis that higher *HBA* copy number is associated with greater risk of kidney disease among Black Americans. Moreover, the protective 2- and 3-copy *HBA* genotypes (i.e., homozygotes and heterozygotes for the 3.7 kb gene deletion) were estimated to prevent approximately 4% of prevalent CKD and 11% of incident ESKD among Black Americans.

*HBA* copy number was associated with both the risk of prevalent CKD at baseline and the incidence of ESKD; *HBA* copy number was not associated with incident reduced eGFR. To be included in the incident reduced eGFR analysis, participants had to undergo a second in-home visit. Participants in the group providing second in-home visit data appeared healthier and of a higher socioeconomic status at baseline than those who did not take part in the second in-home visit. Together, the reduced disease severity, smaller sample size available for analysis (n=3,736), and the stringent pre-specified definition of reduced eGFR (40% decrease), may have reduced the power to detect an effect of *HBA* copy number on this secondary endpoint.

Genetic associations can arise from population structure, i.e., underlying differences in genetic ancestry between those who develop kidney disease and those who do not. To address this, we performed a pre-specified sensitivity analysis that included principal components of ancestry and confirmed the association between *HBA* copy number and prevalent CKD was not due to population structure. We evaluated a pre-specified additive relationship between *HBA* copy number and kidney disease risk because hematologic observations demonstrate a quantitative relationship between *HBA* copy number and alpha globin protein abundance.^45,46^ The association with *HBA* copy number was unlikely due to linkage disequilibrium with sequence variants in nearby genes, given the established relationship between *HBA* copy number and functional gene expression; the rapid decline of linkage disequilibrium away from the *HBA* loci;^47^ and the additive mode of inheritance (i.e., a dose-response relationship between *HBA* copy number and CKD risk) which transcends any single *HBA* allele. Nevertheless, we (i) measured pairwise linkage disequilibrium between the structural variant and SNPs directly and (ii) measured association between each SNP and prevalent CKD. We found no SNPs that could explain the observed association between *HBA* copy number and kidney disease risk. Changes in the level of *HBA* gene expression determined by functional gene copy number remains the most plausible genetic mechanism responsible for the associations with kidney disease risk.

Alpha thalassemia is known to modify the risk of sickle cell disease nephropathy^42,43^; therefore, we considered whether the reduction in *HBA* copy number mitigated the impact of sickle cell trait on kidney disease risk. We found the association between *HBA* copy number and kidney disease risk to be independent of sickle cell trait, and there was no interaction between *HBA* copy number and sickle cell trait on kidney disease outcomes in the pre-specified analysis. A previous study reported that *HBA* genotype modified CKD risk among those with sickle cell trait, but they found no association between *HBA* and CKD in the general population.^44^ The current study differs substantially from the prior report in the following ways: (1) ddPCR was used to quantify *HBA* copy number from 2 to 6; (2) a prespecified log-linear effect of *HBA* copy number was employed; (3) the current study population was more than three times larger, with older individuals and a higher prevalence of CKD; (4) additional established social, demographic, clinical factors were included in the current analysis. Together, the precise estimation of copy number, increased sample size, and additional covariates may have increased the power to detect an association between *HBA* copy number and kidney disease risk in the current study.

There are several potential mechanisms through which *HBA* copy number variation could modify kidney disease risk in a general population of Black Americans. One such mechanism is through decreased erythroid expression of *HBA* leading to anemia, a risk factor for kidney disease. However, *HBA* variants associated with protection against kidney disease tended to be associated with lower, not higher, hemoglobin levels. Moreover, the associations between *HBA* copy number and kidney disease risk remained significant when adjusted for hemoglobin level. We considered whether anemia associated with CKD could have contributed to overestimation of the effect of *HBA* copy number in the prevalent CKD model. In a post-hoc sensitivity analysis omitting hemoglobin, there was no substantial change in the prevalence ratio for CKD (Appendix Table 4 in the Appendix). Red cell microcytosis associated with lower *HBA* copy numbers could potentially confer protection against kidney disease through improved blood rheology;^39^ however, this mechanism does not explain the increased risk associated with higher *HBA* copy numbers (5,6) that have normal MCV. Thus, a clear hematological mechanism explaining the association between *HBA* copy number and kidney disease risk is not apparent.

Alpha globin is expressed not only in red cell precursors, but also in the endothelium of resistance arteries where it regulates nitric oxide signaling.^24^ Genetic deletion of *Hba* or its stabilizing protein *Ahsp* altered small artery vasoreactivity in mice—specifically reducing vasoconstriction to alpha-adrenergic stimuli in a NOS-dependent manner.^24,27^ Pharmacological disruption of the alpha globin/eNOS complex in endothelial cells increased nitric oxide release and led to a fall in blood pressure in both normal and hypertensive mice.^25,40^ We speculate that decreased alpha globin gene expression in individuals with decreased HBA copy number could lead to similar increases in endothelial nitric oxide signaling in human arteries and decreased vasoconstriction in response to sympathetic nervous system activity. This could confer protection against kidney disease in a manner consistent with the protective role of increased nitric oxide signaling in experimental and human clinical studies of kidney injury.^19,15,16,18,41^

This study has strengths and limitations. The REGARDS study is one of the largest available cohorts of Black Americans, and has clearly-defined kidney disease outcome measures and as well as data on social, biomedical, and genetic factors. This allows adjustment for established kidney disease risk factors when estimating risk. We pre-specified the analysis plan to reduce the chance of spurious associations. We employed a robust and quantitative ddPCR method to genotype the insertion/deletion polymorphism and gene triplication in the *HBA* loci. We evaluated genetic risk factors for CKD specific to Black individuals, SCT and high-risk *APOL1* status, and determined that *HBA* copy number was associated with kidney disease independently of these factors. This analysis focused on a Black American population, in which the 3.7 kb gene deletion is common. These findings should not be extrapolated to other populations in which the 3.7 kb deletion is found.

The major genetic variants that determine kidney disease risk among Blacks arose in Africa as mutations conferring resistance to infectious diseases: *HBA* and *HBB* variants (like those that cause alpha thalassemia and sickle cell trait) confer protection against malaria while *APOL1* variants protect against trypanosomiasis, or African sleeping sickness.^48–50^ In contrast to the variant alleles of *HBB* and *APOL1*, which confer protection against infectious diseases at the cost of increased kidney disease risk, the 3.7 kb deletion in *HBA* is associated with protection against both malaria and kidney disease.

In conclusion, we report that higher *HBA* copy number was independently associated with greater CKD prevalence and ESKD incidence after accounting for known clinical, demographic and genetic risk factors in this national longitudinal study of Black Americans. The high frequency of the *HBA* gene deletion found in Black Americans may act to reduce the overall burden of kidney disease in this population.

## Data Availability

The data is maintained by the UAB REGARDS Statistical and Data
Coordinating Center (SDCC).

## ACKNOWLEDGEMENTS

This is an ancillary study supported by cooperative agreement U01 NS041588 co-funded by the National Institute of Neurological Disorders and Stroke (NINDS) and the National Institute on Aging (NIA), National Institutes of Health, Department of Health and Human Service. This research was supported in part by the Divisions of Intramural Research, National Institute of Allergy and Infectious Diseases project AI001150 (A.P.R., H.C.A), National Heart, Lung, and Blood Institute (NHLBI) project HL006196 (A.P.R., Y.Y., H.C.A.). This work was also funded in part by the National Cancer Institute (NCI) Intramural Research Program (C.A.W) and under contract HHSN26120080001E, the National Institute of Diabetes and Digestive and Kidney Diseases (NIDDK) Z01 DK04312 (J.B.K.), and the NHLBI grants K08HL12510 (R.P.N.) and K08HL096841 (N.A.Z.). The content is solely the responsibility of the authors and does not necessarily represent the official views of the NINDS, NIA, NIAID, NCI, NIDDK, or NHLBI. The content of this publication does not necessarily reflect the view or policy of the Department of Health and Human Services, nor does mention of trade names, commercial products or organizations imply endorsement by the government. Some of the data reported here have been supplied by the United States Renal Data System (USRDS). The interpretation and reporting of these data are the responsibility of the author(s) and in no way should be seen as an official policy or interpretation of the U.S. government. The authors thank the investigators, staff, and participants of the REGARDS study for their valuable contributions. A full list of participating REGARDS investigators and institutions can be found at http://www.regardsstudy.org.

