## Appendix for "*HBA* Copy Number and Kidney Disease Risk among Black Americans: a Longitudinal Cohort Study"

[1]Laboratory of Malaria and Vector Research, National Institute of Allergy and Infectious Diseases, National Institutes of Health, Bethesda, MD; [2]Pulmonary Branch, National Heart, Lung, and Blood Institute, Bethesda, MD; [3]Office of Biostatistics Research, National Heart, Lung, and Blood Institute, Bethesda, MD; [4]Division of Blood Diseases and Resources, National Heart, Lung, and Blood Institute, Rockville, MD; [5]Division of Hematology, Department of Medicine, Johns Hopkins University School of Medicine, Baltimore, Maryland; [6]Department of Biostatistics, University of Alabama at Birmingham, Birmingham, Alabama; [7]University of Alabama at Birmingham School of Medicine, Birmingham, Alabama; [8]Department of Medicine, Larner College of Medicine at the University of Vermont, Burlington, Vermont; [9]Department of Pathology & Laboratory Medicine, Larner College of Medicine at the University of Vermont, Burlington, Vermont; [10]Basic Research Program, National Cancer Institute, Frederick National Laboratory for Cancer Research, Frederick, Maryland; [11]Kidney Diseases Branch, National Institute of Diabetes and Digestive and Kidney Diseases, National Institutes of Health, Bethesda, Maryland; [12]Division of Biomedical Informatics and Personalized Medicine, Department of Medicine, University of Colorado, Denver, CO; [13]Department of Epidemiology, University of Alabama at Birmingham, Birmingham, Alabama; [14]Department of Medicine, University of Alabama at Birmingham, Birmingham, Alabama

**I. Supplemental Figure and Tables**......……………………………………………………...3

Appendix Figure 1. Comparison of prevalence ratios for *HBA*, *APLO1*, and *HBB* genetic risk factors for CKD..........................................................................................3

Appendix Figure 2. Linkage disequilibrium and association analysis of sequence variants in the first 1 Mb of chromosome 16 flanking *HBA1* and *HBA2*...................................................................................................................4

Appendix Table 1. Association of *HBA* copy number with prevalent CKD, incident reduced eGFR, and incident ESKD – fully adjusted models including ten principal components of ancestry………………………………………………………………….…6

Appendix Table 2. Pre-specified test for interaction between *HBA* copy number and SCT on the outcomes of prevalent CKD, incident reduced eGFR, and incident ESKD in fully adjusted models………………………….…………...............……………….....................…8

Appendix Table 3. Pre-specified tests for interaction between each of Age, Sex, Hypertension, or APOL1 and *HBA* on the outcome of prevalent CKD in fully adjusted models…………………………………………………………………………...……...9

Appendix Table 4. Post-hoc sensitivity analysis of the association of *HBA* copy number with prevalent CKD when hemoglobin is omitted from the model………...........................................................................................................10

**II. Estimation of population preventable fraction of *HBA* copy number on kidney disease**………………………………………………………………………….…..12

**III. Additional Methods**………………………………………….....……………….…16

a. *HBA* Genotyping Methods……………………………………………………….…16
b. Multiple Imputation Procedure……………………………….…………………….…16
c. Diagnostic Modeling Description…………………………………………….…….…17
d. Assessment of the Missing at Random Assumption……………………………….…18

**IV. Supplemental References**……………………………………………………….……20

### I. Appendix Figure and Tables

**Appendix Figure 1. Comparison of prevalence ratios for *HBA*, *APLO1*, and *HBB* genetic risk factors for CKD**

| Locus | PR | 95% CI |
|---|---|---|
| ***HBA*** | | |
| 2 copies | 0.77 | (0.68 − 0.86) |
| 3 copies | 0.88 | (0.83 − 0.93) |
| 4 copies (ref) | 1 | − |
| 5−6 copies | 1.14 | (1.07 − 1.21) |
| ***APOL1*** | | |
| High risk | 1.16 | (1.05 − 1.28) |
| ***HBB*** | | |
| Sickle cell trait | 1.43 | (1.29 − 1.59) |

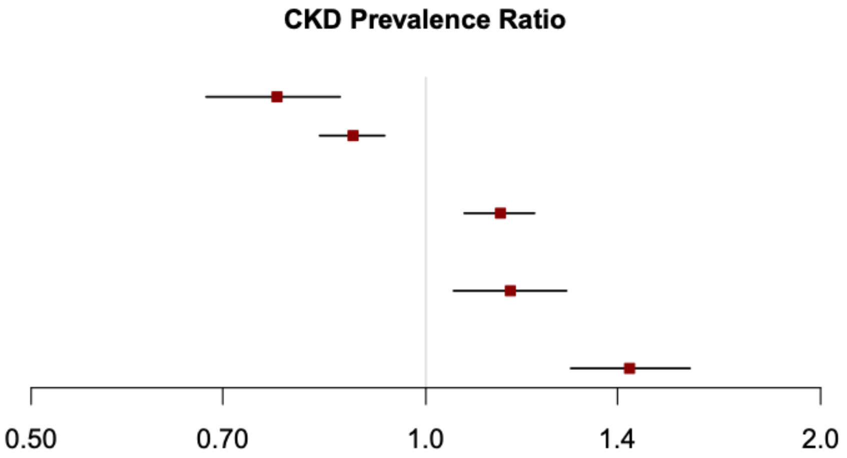


*HBA*= alpha globin gene; *APOL1*= apolipoprotein-1; *HBB*= hemoglobin beta-sickle cell trait; CKD= chronic kidney disease; PR= prevalence ratio

Supplemental Figure 1 Legend. Prevalence ratio of chronic kidney disease by alpha globin *HBA* copy number, *APOL1*, and sickle cell trait *HBB-SCT* –adjusted analysis. Prevalence ratios of genetic risk factors for chronic kidney disease represented on a log scale. Depicted prevalence ratios for each *HBA* copy number were calculated based on the reported modified Poisson multivariable regression model adjusting for 13 risk factors for chronic kidney disease.

**Appendix Figure 2. Linkage disequilibrium and association analysis of sequence variants in the first 1 Mb of chromosome 16 flanking *HBA1* and *HBA2*.**

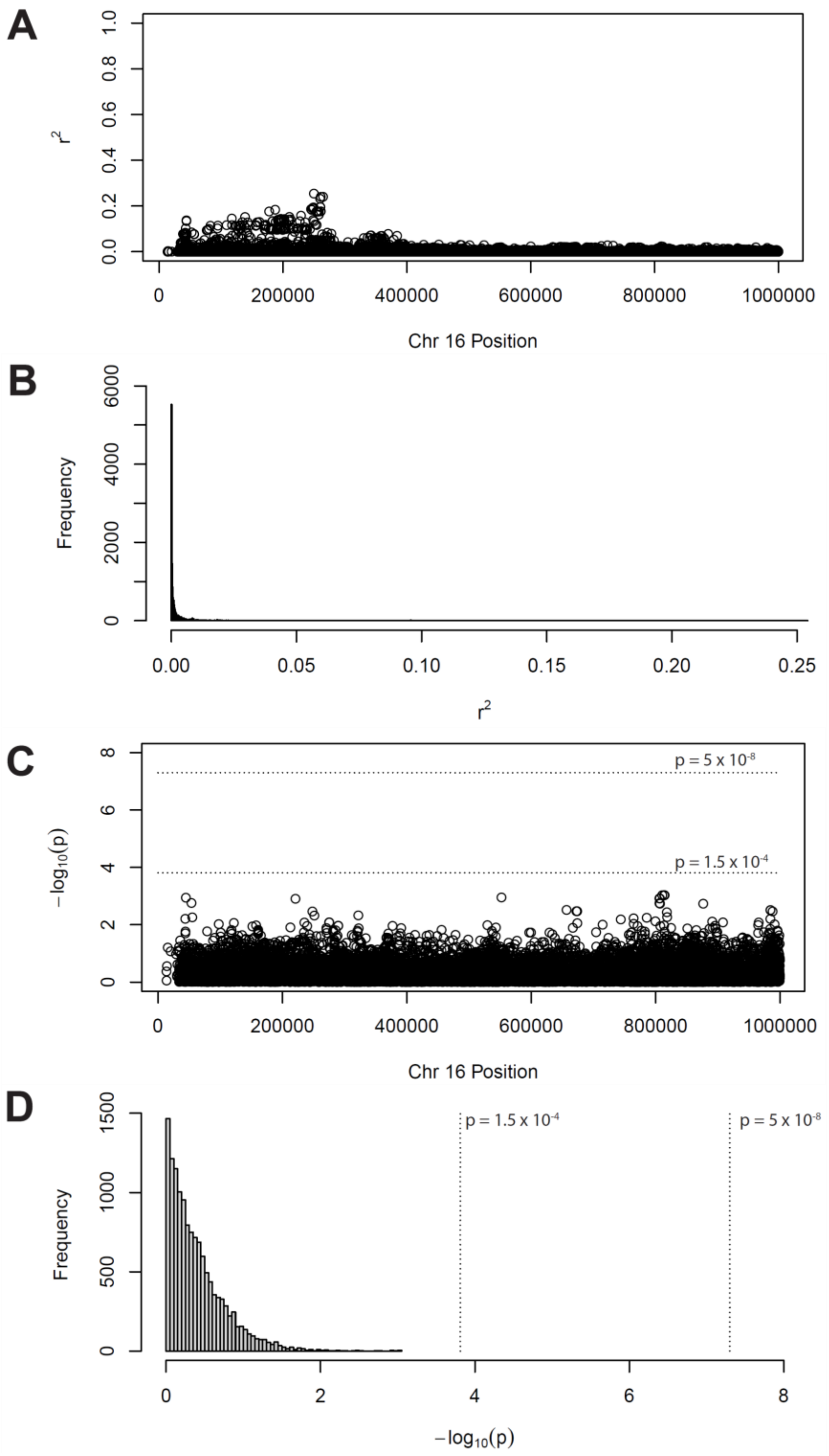

**Appendix Figure 2. A**, Pairwise linkage disequilibrium ($r^2$) between the -3.7 kb structural variant and SNPs genotyped on the Infinium Expanded Multi-Ethnic Genotyping Array in 8,841 study participants. The deletion is found at position 173619-177403. **B**, Histogram showing the frequency distribution of $r^2$ values. **C**, Association between each SNP and prevalent CKD using a model that includes the age, sex, and the first 4 principal components of ancestry. Horizontal lines indicate the p-value significance thresholds for genome-wide significance ($5 \times 10^{-8}$) and for a 1 Mb region ($1.5 \times 10^{-4}$). **D**, Histogram showing the frequency distribution of association tests for all SNPs in the 1 Mb region of chromosome 16.

**Appendix Table 1. Association of *HBA* copy number with prevalent CKD, incident reduced eGFR, and incident ESKD – fully adjusted models including ten principal components of ancestry.**

| | **Prevalent chronic kidney disease*** Modified Poisson (n=7,641) | | **Incident reduced eGFR**[†] Modified Poisson (n=2,540) | | **Incident end-stage kidney disease**[‡] Cox proportional hazards (n=7,509) | |
|---|---|---|---|---|---|---|
| | **PR** | **CI** | **RR** | **CI** | **HR** | **CI** |
| ***HBA* copy number, per gene copy** | 1.16 | (1.08 - 1.23) | 1.03 | (0.88 - 1.20) | 1.28 | (1.01 - 1.63) |
| **PCA**[‖] | | | | | | |
| **PCA 1** | 1.02 | (0.98 - 1.06) | 0.99 | (0.91 - 1.09) | 0.94 | (0.80 - 1.09) |
| **PCA 2** | 0.98 | (0.95 - 1.02) | 1.04 | (0.91 - 1.09) | 1.20 | (1.05 - 1.38) |
| **PCA 3** | 1.01 | (0.97 - 1.04) | 0.95 | (0.87 - 1.05) | 0.98 | (0.85 - 1.12) |
| **PCA 4** | 1.02 | (0.98 - 1.05) | 1.00 | (0.92 - 1.09) | 0.94 | (0.83 - 1.07) |
| **PCA 5** | 1.02 | (0.98 - 1.06) | 0.97 | (0.89 - 1.07) | 0.94 | (0.82 - 1.07) |
| **PCA 6** | 1.00 | (0.97 - 1.04) | 0.99 | (0.91 - 1.09) | 0.84 | (0.73 - 0.96) |
| **PCA 7** | 0.96 | (0.93 - 1.00) | 1.02 | (0.94 - 1.12) | 0.97 | (0.85 - 1.11) |
| **PCA 8** | 1.01 | (0.97 - 1.05) | 1.03 | (0.94 - 1.13) | 1.03 | (0.90 - 1.18) |
| **PCA 9** | 0.98 | (0.94 - 1.01) | 0.98 | (0.90 - 1.07) | 1.04 | (0.91 - 1.18) |
| **PCA 10** | 1.01 | (0.98 - 1.05) | 0.94 | (0.86 - 1.02) | 0.94 | (0.83 - 1.08) |

*HBA*= alpha globin gene; CKD= chronic kidney disease; eGFR= estimated glomerular filtration rate; ESKD= end-stage kidney disease; PR= prevalence ratio; CI= 95% confidence interval; RR= relative risk; HR = hazard ratio; PCA= principal components of ancestry.

[*]Prevalent chronic kidney disease was defined by eGFR < 60mL/min/1.73m$^2$ or urine albumin to creatinine ratio ≥ 30mg/g. [†]Incident reduced eGFR was defined by an eGFR < 60mL/min at the

follow-up in-home visit and greater than 40% decline in eGFR from baseline, among those who had eGFR ≥ 60 mL/min at baseline. [‡]Incident end-stage kidney disease (ESKD) was identified by linkage to the United States Renal Data System data through June 30, 2014. [§]P values were calculated using either modified Poisson or Cox proportional hazards multivariable regression models, as noted, employing a linear effect of *HBA* allele count; [||] 8,841 participants had GWAS data available for PCA analysis. The prevalent CKD model had n=7,641 subjects with data available for the PCA analysis, the incident reduced eGFR model had n=2,540 subjects, and the incident ESKD model had n=7,509 subjects. Multiple imputations were performed for other missing data.  The following variables were included in the model but are not displayed in this table:  Sickle cell trait, *APOL1* high-risk status, hemoglobin, age, sex, body mass index, hypertension, diabetes mellitus, smoking status, medically insured, region, education level, income.

**Appendix Table 2. Pre-specified test for interaction between *HBA* copy number and SCT on the outcomes of prevalent CKD, incident reduced eGFR, and incident ESKD in fully adjusted models.**

| | Prevalent chronic kidney disease* Modified Poisson (n=9,918) | | | Incident reduced eGFR† Modified Poisson (n=3,736) | | | Incident end-stage kidney disease‡ Cox proportional hazards (n=9,707) | | |
|---|---|---|---|---|---|---|---|---|---|
| | **PR** | **CI** | ***P value*§** | **RR** | **CI** | ***P value*§** | **HR** | **CI** | ***P value*§** |
| **SCT*HBA** | 1.13 | (0.93 - 1.38) | 0.23 | 0.99 | (0.62 - 1.59) | 0.97 | 1.40 | (0.65 - 2.99) | 0.39 |

SCT= sickle cell trait; *HBA*= alpha globin gene; CKD= chronic kidney disease; eGFR= estimated glomerular filtration rate; ESKD= end-stage kidney disease; PR= prevalence ratio; CI= 95% confidence interval; RR= relative risk; HR= hazard ratio

*Chronic kidney disease was defined by estimated glomerular filtration rate [GFR] <60mL/min/1.73m$^2$ or urine albumin to creatinine ratio $\geq$30mg/g. †Incident reduced eGFR was defined by an eGFR < 60mL/min at the follow-up in-home visit and greater than 40% decline in eGFR from baseline, among those who had eGFR ≥ 60 mL/min at baseline. ‡Incident end-stage kidney disease (ESKD) was identified by linkage to the United States Renal Data System data through June 30, 2014. §P values were calculated using either modified Poisson or Cox proportional hazards multivariable regression models, as noted, employing a monotonic effect of *HBA* allele count. The prevalent CKD model had n=9,918 subjects, the incident reduced eGFR model had n=3,736 subjects, and the incident ESKD model had n=9,707 subjects. Multiple imputations were performed for missing data. The following variables were included in the model but are not displayed in this table: *HBA* copy number, sickle cell trait, *APOL1* high-risk status, hemoglobin, age, sex, body mass index, hypertension, diabetes mellitus, smoking status, medically insured, region, education level, income.

**Appendix Table 3. Pre-specified tests for interaction between each of Age, Sex, Hypertension, or *APOL1* and *HBA* on the outcome of prevalent CKD in fully adjusted models.**

| Separate fully adjusted models with interaction terms individually added | Prevalent chronic kidney disease† Modified Poisson (n=9,918) | | |
|---|---|---|---|
| | **PR** | **CI** | ***P value*‡** |
| **Age*HBA** | 1.00 | (0.99 - 1.01) | 0.72 |
| **Female Sex*HBA** | 0.93 | (0.83 - 1.04) | 0.21 |
| **Hypertension*HBA** | 0.99 | (0.79 - 1.25) | 0.96 |
| ***APOL1* high-risk genotype*HBA** | 0.97 | (0.83 - 1.13) | 0.66 |

*APOL1*= apolipoprotein-L1; *HBA*= alpha globin; CKD= chronic kidney disease; PR= prevalence ratio; CI= 95% confidence interval.

†Prevalent chronic kidney disease was defined by estimated glomerular filtration rate <60mL/min and/or urine albumin creatinine ratio $\geq$ 30 mg/g. ‡Each interaction term was added separately to the main prevalent chronic kidney disease model. P values were calculated using modified Poisson multivariable regression models employing a linear effect of *HBA* allele count on the log of the relative risk with all clinical and demographic covariates utilized in the main prevalent chronic kidney disease model. The following variables were included in the model but not displayed in this table: *HBA* copy number, sickle cell trait, *APOL1* high-risk status, hemoglobin, age, sex, hypertension, diabetes mellitus, body mass index, smoking status, medically insured, region, education level, income.

**Appendix Table 4. Post-hoc sensitivity analysis of the association of *HBA* copy number with prevalent CKD when hemoglobin is omitted from the model.**

| | Prevalent chronic kidney disease* (*n*=9,918) Modified Poisson | |
|---|---|---|
| | **PR** | **CI** |
| ***HBA* copy number, per gene copy** | 1.10 | (1.03 - 1.16) |
| **Sickle cell trait** | 1.46 | (1.32 - 1.62) |
| ***APOL1* high-risk** | 1.18 | (1.07 - 1.29) |
| **Age, per year** | 1.03 | (1.03 - 1.04) |
| **Female sex** | 0.88 | (0.82 - 0.95) |
| **Body mass index**[†] | 1.09 | (1.05 - 1.12) |
| **Hypertension** | 1.99 | (1.74 - 2.26) |
| **Diabetes mellitus** | 1.72 | (1.61 - 1.84) |
| **Smoking status** | | |
| Never (ref) | - | - |
| Past | 0.97 | (0.90 - 1.04) |
| Present | 1.22 | (1.11 - 1.35) |
| **Medically Insured** | 0.93 | (0.82 - 1.05) |
| **Region** | | |
| Non-Belt (ref) | - | - |
| Belt | 0.97 | (0.90 - 1.04) |
| Buckle | 1.02 | (0.93 - 1.11) |

| | | |
|---|---|---|
| **Education level** | | |
| < HS Grad (ref) | - | - |
| HS Grad | 0.93 | (0.85 - 1.02) |
| Some College | 0.98 | (0.89 - 1.08) |
| ≥ College Grad | 0.93 | (0.83 - 1.03) |
| **Income** | | |
| < $20K (ref) | - | - |
| $20K - $34K | 0.97 | (0.89 - 1.05) |
| $35K - $74K | 0.90 | (0.81 - 1.01) |
| ≥ $75K | 0.67 | (0.56 - 0.81) |

*HBA*= alpha globin gene; CKD= chronic kidney disease; PR= prevalence ratio; CI= 95% confidence interval; APOL1= apolipoprotein-L1; (ref) indicates reference category; K= thousand.

*Prevalent CKD was defined by a urine albumin to creatinine ratio ≥ 30 mg/g or an estimated glomerular filtration rate (eGFR) < 60 mL/min/1.73m2 at baseline. Analysis performed using modified Poisson multivariable regression model employing a linear effect of *HBA* allele count on the log of the prevalence ratio. †Body mass index scaled by standard deviation. All variables shown in table were included in the multivariable model. Multiple imputations were performed for missing data. The number of subjects available for this analysis was 9,918.

### II. Estimation of population preventable fraction of *HBA* copy number on kidney disease

The observed population had the following allele frequencies:

| *HBA* copy number | 2 | 3 | 4 | 5 or 6 |
|---|---|---|---|---|
| Prevalence, % | 4.0 | 27.7 | 67.3 | 1.0 |

The population preventable fraction measures the degree to which the elimination of a beneficial risk factors in a population increases the corresponding population prevalence or risk of disease. The population attributable fraction measures the degree to which elimination of a detrimental risk factor decreases the population prevalence or risk.

#### Prevalent Chronic Kidney Disease

To estimate the fraction of disease prevalence that is prevented by reductions in *HBA* copy number, we first dichotomized the *HBA* copy number into two classes: (2, 3) versus (4, 5, 6) and asked to what extent is chronic kidney disease (CKD) prevalence decreased by the (2, 3) allele class.

We used a fully-adjusted Poisson regression model to estimate the risk associated with the (4, 5, 6) class versus the (2, 3) class:

| **Variables** | **PR** | **CI** | **P value** |
|---|---|---|---|
| *HBA* copy number (4, 5, 6) | 1.14 | (1.06 - 1.23) | 0.0002 |
| Sickle cell trait | 1.44 | (1.30 - 1.59) | < 0.0001 |
| *APOL1* high risk | 1.17 | (1.07 - 1.29) | 0.0006 |
| Hemoglobin, per 1g/dL | 0.90 | (0.87 - 0.93) | < 0.0001 |
| Age, per year | 1.03 | (1.03 - 1.04) | < 0.0001 |
| Female sex | 0.77 | (0.71 - 0.84) | < 0.0001 |
| Body mass index | 1.09 | (1.05 - 1.12) | < 0.0001 |
| Hypertension | 1.96 | (1.72 - 2.23) | < 0.0001 |
| Diabetes mellitus | 1.64 | (1.53 - 1.76) | < 0.0001 |
| Smoking, past | 0.97 | (0.91 - 1.05) | 0.49 |

| | | | |
|---|---|---|---|
| Smoking, present | 1.29 | (1.17 - 1.42) | < 0.0001 |
| Medically insured | 0.92 | (0.81 - 1.04) | 0.19 |
| Region belt | 0.96 | (0.89 - 1.04) | 0.33 |
| Region buckle | 1.00 | (0.91 - 1.09) | 0.96 |
| Education level high school graduate | 0.94 | (0.86 - 1.03) | 0.21 |
| Education level some college | 0.99 | (0.90 - 1.10) | 0.92 |
| Education level college graduate or more | 0.94 | (0.84 - 1.05) | 0.29 |
| Income $20k-$34k | 0.97 | (0.89 - 1.06) | 0.49 |
| Income $35k-$74k | 0.91 | (0.81 - 1.01) | 0.07 |
| Income $75k and above | 0.69 | (0.57 - 0.83) | < 0.0001 |

Using methods based on Greenland and Drescher adapted to modified Poisson regression and multiple imputations we then estimated the population preventable fraction (PPF) as the increased prevalence of CKD that would exist in an alternative REGARDS population that has only the (4, 5, 6) allele class:

$$\text{PPF} = \frac{\text{Risk}_{\text{Alternative Population}} - \text{Risk}_{\text{Population Observed}}}{\text{Risk}_{\text{Population Observed}}}$$

The prevalence of CKD in the alternative REGARDS population would increase by 4.3% (95% CI 2.1 - 6.5).

For comparison, a similar calculation of population attributable fraction (PAF) was performed for sickle cell trait (PAF was computed instead of the population preventable fraction because sickle cell trait is harmful, rather than beneficial). The prevalence of CKD in an alternative REGARDS population that does not carry sickle cell trait would decrease by 3.1 % (95% CI 2.1 - 4.2).

In conclusion, a reduction in *HBA* copy number explains a non-zero fraction of CKD risk that is similar in size to that attributable to sickle cell trait. While the population preventable fraction point estimate for *HBA* deletions is greater than the population attributable fraction for sickle cell trait (4.5% vs 3.1%), the 95% confidence intervals for these estimates overlap.

**Incident End-Stage Chronic Kidney Disease**

To estimate the fraction of disease risk that is attributable to deletions in *HBA* copy number, we first dichotomized the *HBA* copy number into two classes: (2, 3) versus (4, 5, 6).

We used a fully-adjusted Cox proportional hazards model to estimate the risk associated with the (4, 5, 6) class versus the (2, 3) class:

| Variables | HR | CI | P value |
|---|---|---|---|
| *HBA* copy number (4, 5, 6) | 1.42 | (1.06 - 1.89) | 0.02 |
| Sickle cell trait | 2.15 | (1.47 - 3.13) | < 0.0001 |
| *APOL1* high risk | 1.83 | (1.32 - 2.53) | 0.0003 |
| Hemoglobin, per 1g/dL | 0.57 | (0.52 - 0.63) | < 0.0001 |
| Age, per year | 1.00 | (0.99 - 1.02) | 0.75 |
| Female sex | 0.38 | (0.28 - 0.52) | < 0.0001 |
| Body mass index | 1.08 | (0.94 - 1.24) | 0.25 |
| Hypertension | 7.50 | (3.04 - 18.50) | < 0.0001 |
| Diabetes mellitus | 3.59 | (2.68 - 4.81) | < 0.0001 |
| Smoking, past | 1.32 | (0.98 - 1.78) | 0.07 |
| Smoking, present | 2.15 | (1.46 - 3.18) | 0.0001 |
| Medically insured | 1.04 | (0.64 - 1.70) | 0.87 |
| Region belt | 0.84 | (0.62 - 1.14) | 0.27 |
| Region buckle | 0.86 | (0.59 - 1.25) | 0.44 |
| Education level high school graduate | 1.49 | (1.02 - 2.18) | 0.04 |
| Education level some college | 1.19 | (0.78 - 1.82) | 0.41 |
| Education level college graduate or more | 1.81 | (1.16 - 2.83) | 0.009 |
| Income $20k-$34k | 0.89 | (0.65 - 1.23) | 0.48 |

| | | | |
|---|---|---|---|
| Income \$35k-\$74k | 0.46 | (0.30 - 0.71) | 0.0005 |
| Income \$75k and above | 0.24 | (0.10 - 0.58) | 0.002 |

Finally, applying methods of Zetterqvist et al. in a setting with multiple imputations we estimated the population preventable fraction (PPF) for the 2,3 class as the increased risk of incident end-stage kidney disease (ESKD) at follow-up time *t* that would exist in an alternative REGARDS population that had only the (4, 5, 6) allele class:

$$\mathrm{PPF} = \frac{\mathrm{Prob(ESRD\ by\ time\ t)}_{\text{Alternative Population}} - \mathrm{Prob(ESRD\ by\ time\ t)}_{\text{Population Observed}}}{\mathrm{Prob(ESRD\ by\ time\ t)}_{\text{Population Observed}}}$$

The risk of incident ESKD in the alternative REGARDS population would increase by 11.4%, (95% CI 2.2 - 20.6). This was estimated at the median follow-up time of 10.1 years, and calculations at the 25th and 75th percentile of follow-up time gave similar results.

For comparison, a similar calculation was performed for sickle cell trait. The risk of incident ESKD at 10.1 years of follow-up in an alternative REGARDS population that does not carry sickle cell trait would decrease by 7.3% (95% CI 2.8 - 11.8).

In conclusion, a reduction in alpha globin gene copy number explains a non-zero fraction of CKD risk that is similar in size to that attributable to sickle cell trait. While the PPF point estimate for *HBA* deletions is greater than the PAF for sickle cell trait (11.4% vs 7.3%), the 95% confidence intervals for these estimates overlap.

### III. Additional Methods

#### a. *HBA* Genotyping Methods

Two-dimensional clusters of droplet counts for target and reference genes were manually gated using Quantasoft (Bio-Rad) per the manufacturer's protocols. Droplet counts, copy number variant (CNV) values, and 95% CIs for CNV were extracted, visualized, and genotype was assigned using custom scripts in the R computing environment without user intervention. A subset of samples was validated against an independent approach employing multiple ligation-dependent probe amplification (MLPA) performed at the Mayo Clinic Laboratory, with 100% concordance. Inter-day variation of our assay was determined by performing the assay on two different days on 672 samples; quantitative copy number varied by less than 1% between days. Reference samples of known genotype were run as positive controls and reaction wells with water instead of DNA were run as negative controls each day.

#### b. Multiple Imputation Procedure

Multiple imputation methods were used in the multivariable analyses. Data on the degree of missingness for the outcome of prevalent chronic kidney disease are described in the footnote to Table 1 in the manuscript. Missing patterns for incident reduced eGFR and incident end-stage kidney disease were similar to those detailed for the prevalent chronic kidney disease analysis. The R package “mice” Version 3.6.0 was used to create and analyze the resulting imputations.[1] Each analysis presented is based upon 20 imputations (each developed using 30 Markov Chain based iterations) and the final model coefficients and their standard errors were derived using Rubin’s method for pooling results across imputations.[2] The variables used in the imputation procedure were those used in the corresponding regression model and sometimes augmented with additional variables (e.g. systolic and diastolic blood pressure for determining hypertension, and albumin creatinine ratio and eGFR for CKD prevalence). The imputations evolution over 30 iterations was examined visually for convergence and mixing. Further, the distributions of complete and imputed values were visually examined for aberrations.

**c. Diagnostic Modeling Description**

Our modeling was prespecified in our analytic plan, as described. We performed diagnostic investigation of the Poisson models (for CKD prevalence and CKD incidence) and Cox models (for ESKD incidence) in order to guide sensitivity analyses.

For the Poisson models involving CKD prevalence and CKD incidence the R function "glm" and R package "sandwich" were used. Residuals were examined for evidence of poor fitting as evidenced by correlation between residuals and predictors of fitted values. Testing of Pearson residuals indicated that age and hemoglobin were perhaps inadequately modeled as having linear relationships on the log of the risk for CKD prevalence. Consequently, we extended our main model to include quadratic terms for age and hemoglobin. These additional terms had significant p-values but did not change the results for allele count or sickle cell trait in any meaningful way (point estimates of the risk ratio changed from 1.14 to 1.13 and 1.43 to 1.42 and both p-values remained $< 0.0001$). Conducting a similar analysis for incident reduced eGFR suggested the effect of age on log of risk was not adequately captured through a linear relationship and a quadratic term was added. The estimated effect for allele count was not qualitatively changed and remained non-significant (risk ratio estimate of 1.02, $p = 0.73$). For both CKD prevalence and reduced eGFR incidence Pearson residuals were not significantly correlated with allele count.

For the model of ESKD incidence using Cox proportional hazard techniques, Schoenfeld residuals were examined over follow-up time to detect violations of proportional hazards assumptions for the covariates in the analysis of ESKD outcomes. There was some evidence of proportional hazards violation for the baseline age covariate ($p < 0.05$ for a chi-square test of non-proportional hazards in the majority of imputed datasets), however including a time by age interaction to address non-proportionality did not qualitatively alter the estimated effects for the other covariates – in particular the p-values for *HBA*, sickle cell trait, and hemoglobin remained significant and of similar magnitude as without the interaction.

**d. Assessment of the Missing at Random Assumption**

Missingness was generally rare with the exception of hemoglobin (32%), self-reported income (12%) and hypertension (4%). Hemoglobin is missing primarily because it was not initially collected for approximately the first 8000 of the REGARDS 30239 participants (all races combined). Given the administrative nature of the missing data an assumption of hemoglobin missing at random (i.e., the probability of missing depends on observed information rather than the underlying missing hemoglobin value) multiple imputation was used.

Income data reported as missing reflect refusal to provide information. These self-reported incomes might not be missing at random as the refusals might more likely coincide with higher or lower than average incomes. As a sensitivity analysis we first imputed the annual income category (either "less than $20k", "$20k-$34k", "$35k-$74k", or "$75k and above") using the multiple imputation algorithm and then moved the imputed category values one level higher if they were not already in the highest category. For example, if a person had an original imputed value of $20k-$34k then in this sensitivity analysis they would now have a value of $35k-$74k. This corresponds to people refusing to answer having higher incomes than predicted. While the education and income coefficients change marginally in the resulting analysis, the remaining coefficients and p-values are essentially unchanged from those presented in the incident ESKD analysis (the HR estimate and p-value for the HBA copy number are 1.27 and 0.04 - essentially unchanged from 1.28 and 0.04). The results when lowering (instead of raising) the imputed income category are qualitatively similar with the resulting HR estimate and p-value for HBA copy number 1.28 and 0.04. These results suggest that using a missing at random assumption for income is reasonable.

A similar analysis was performed with respect to missing values for baseline hypertension (379 of 9,707 individuals in the analysis of incident ESKD). When all 379 were imputed as non-hypertensive the results changed very little for all covariates except hypertension (and this changed only modestly). The HR estimate and p-value for HBA copy number remain 1.28 and 0.04. When all 379 were imputed as hypertensive the results again changed hardly at all (HR estimate and p-value for HBA copy number remain 1.28 and 0.04), again suggesting that using a missing at random assumption for hypertension does not likely lead to misleading estimates for any of the covariates.

These sensitivity analyses were conducted for the time to ESRD analysis as the primary results are close to the 0.05 significance threshold for the HBA copy number effect. Given the strength of the association for the analysis of prevalent CKD we again expect the use of multiple imputation with a missing at random assumption does not generate misleading results because of violations of this assumption. Similarly, we do not expect the non-significant findings for HBA copy number on incident CKD to be qualitatively changed by plausible modifications to the missing at random assumption.
